# Positive-control Mendelian randomization highlights power constraints in disease-mortality GWAS

**DOI:** 10.64898/2026.05.29.26354472

**Authors:** Chen-Yang Su, Guillaume Butler-Laporte

## Abstract

Yang et al. recently published a systematic comparison of genetic effects on disease susceptibility and disease-specific mortality across nine common diseases and seven biobanks, concluding that susceptibility and survival architectures overlap only modestly. This is an important resource, but we argue that the current mortality genome-wide association studies (GWAS) require explicit power calibration before limited overlap can be interpreted biologically. Using two-sample Mendelian randomization (MR) with positive-control exposures, we show that even a well-powered positive control, body mass index (BMI), instrumented by 855 genome-wide-significant variants, produces a clearly detectable effect for heart failure (HF) mortality, with only weaker evidence for chronic kidney disease (CKD) mortality. However, when BMI instruments were stratified into quartiles by exposure-association strength, the heart failure association remained nominally significant only in the two strongest quartiles and was not significant in the two weakest quartiles. Further, using household income as a weakly instrumented socio-economic contrast has insufficient power to detect moderate effects on any disease mortality outcome. These analyses indicate that current disease mortality GWAS may be insufficiently powered to detect shared effects. In contrast, the same BMI instrument set produced large and directionally coherent effects when applied to case-control GWAS of the matched six diseases, with the HF and prostate cancer associations preserved under a within-family BMI sensitivity analysis, and nominal support for CKD. The HF mortality association was also preserved in a within-family BMI sensitivity analysis. Similarly, genetically proxied household income was associated with HF risk in the case-control GWAS despite null associations with disease-specific mortality, consistent with limited power in the mortality GWAS. These findings indicate that the limited BMI-mortality evidence across several outcomes is unlikely to reflect a weak BMI instrument or dynastic artefacts alone and instead supports limited effective power in current disease-mortality GWAS.

## Introduction

Disease mortality GWAS are inherently difficult to power. Cases must be diagnosed, followed and ascertained for a progression endpoint, substantially shrinking effective sample sizes relative to incidence GWAS. Yang et al. previously released the largest systematic comparison to date of disease-risk and disease-specific-mortality GWAS across nine common diseases, concluding that the two architectures overlap only modestly^1^. This represents an important achievement. However, inspection of the mortality summary statistics shows that, for the six diseases with full data (Alzheimer’s disease [AD], chronic kidney disease [CKD], colorectal cancer, heart failure [HF], prostate cancer and stroke), no single nucleotide polymorphism (SNP) reached *P* < 5×10^−8^ (minimum *P* = 1.04×10^−6^, stroke). This in itself is not conclusive evidence of low power, because it can also reflect small effect sizes or higher polygenicity. However, it is a warning sign.

Here we use a different approach to measure the problem by two-sample Mendelian randomization (MR) with positive-control exposures chosen to span statistical power and biological plausibility for mortality. If a GWAS is well-powered, then we would expect to find significant MR associations. We used BMI from Yengo et al.^2^ (N = 681,275) as a highly polygenic positive control with prior MR^3^, observational^4^ and randomised-trial evidence^5^ of causal effects on cardiometabolic mortality, and household income from Kweon et al.^6^ as a weakly instrumented socio-economic exposure often associated with disease risk. To assess whether population-based BMI associations were influenced by demographic, assortative-mating or dynastic effects, we additionally repeated the BMI-mortality analyses using within-family BMI.

## Methods

We included the six mortality GWAS with full effect sizes and standard errors in Yang et al.^1^, as these are required for MR analyses. For each exposure we retained autosomal variants with minor allele frequency (MAF) > 0.01, excluded the major histocompatibility complex region, and clumped at *P* < 5×10^−8^, and *r*^*2*^ < 0.001. Where possible, we used SNP proxies for missing instruments in the outcome GWAS (*r*^*2*^ ≥ 0.8). The primary estimator was inverse-variance-weighted (IVW), with other sensitivity analyses reported in **Supplementary Table S1**.

To test the dependence of detected effects on instrument strength, we stratified independent BMI variants into quartiles (Q1–Q4) by BMI exposure-association *P*-value, with Q1 containing the strongest BMI-associated instruments and Q4 containing the weakest and repeated the MR within each stratum. We computed, for each exposure-outcome pair, the minimum odds ratio (OR) detectable at two-sided α = 0.05 and 80% power,

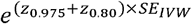

and the power to detect OR values of 1.10 and 1.20 given the observed *SE*_*IVW*_. All analyses used TwoSampleMR v0.5.6 and R v4.1.2; full per-estimator results are in **Supplementary Table S1** and per-pair power in **Supplementary Table S2**.

To validate our positive controls, we additionally ran two-sample MR of each exposure against case-control GWAS of the same six diseases: AD^7^, CKD^8^, colorectal cancer^9^, HF^10^, prostate cancer^11^, and ischaemic stroke^12^. As a sensitivity analysis, we repeated the BMI analysis using within-family BMI^13^ (N = 140,883) as exposure. This conditions on parental genotype and therefore largely removes demographic, assortative-mating and dynastic confounding, at the cost of lower statistical power (9–10 within-family BMI instruments per outcome, including proxies).

## Results

With the full BMI instrument set (up to 855 variants), we observed a clear MR association with HF mortality (OR 1.16 per 1-SD increase in BMI, 95% confidence interval (CI) 1.07–1.26, *P* = 3.5×10^−4^); sensitivity methods were concordant (**Supplementary Table S1**). A less significant effect was observed for CKD mortality (OR 1.21, 95% CI 1.01–1.45, *P* = 0.04). We did not detect associations with AD, prostate cancer, colorectal cancer or stroke mortality (*P* > 0.6; **Figure 1a**). Household-income estimates were imprecise across all six mortality outcomes (**Figure 1a**). Down-sampling the BMI instruments by exposure *P*-value quartile showed that the HF signal became unstable in smaller, weaker strata (**Figure 1b**). Consistent with attenuation after instrument down-sampling, the BMI-HF association remained nominally significant in Q1 (218 variants; OR 1.13, *P* = 0.039) and Q2 (214 variants; OR 1.25, *P* = 0.015), but not in Q3 or Q4 (*P* = 0.14 and 0.30, respectively).

**Figure 1.**
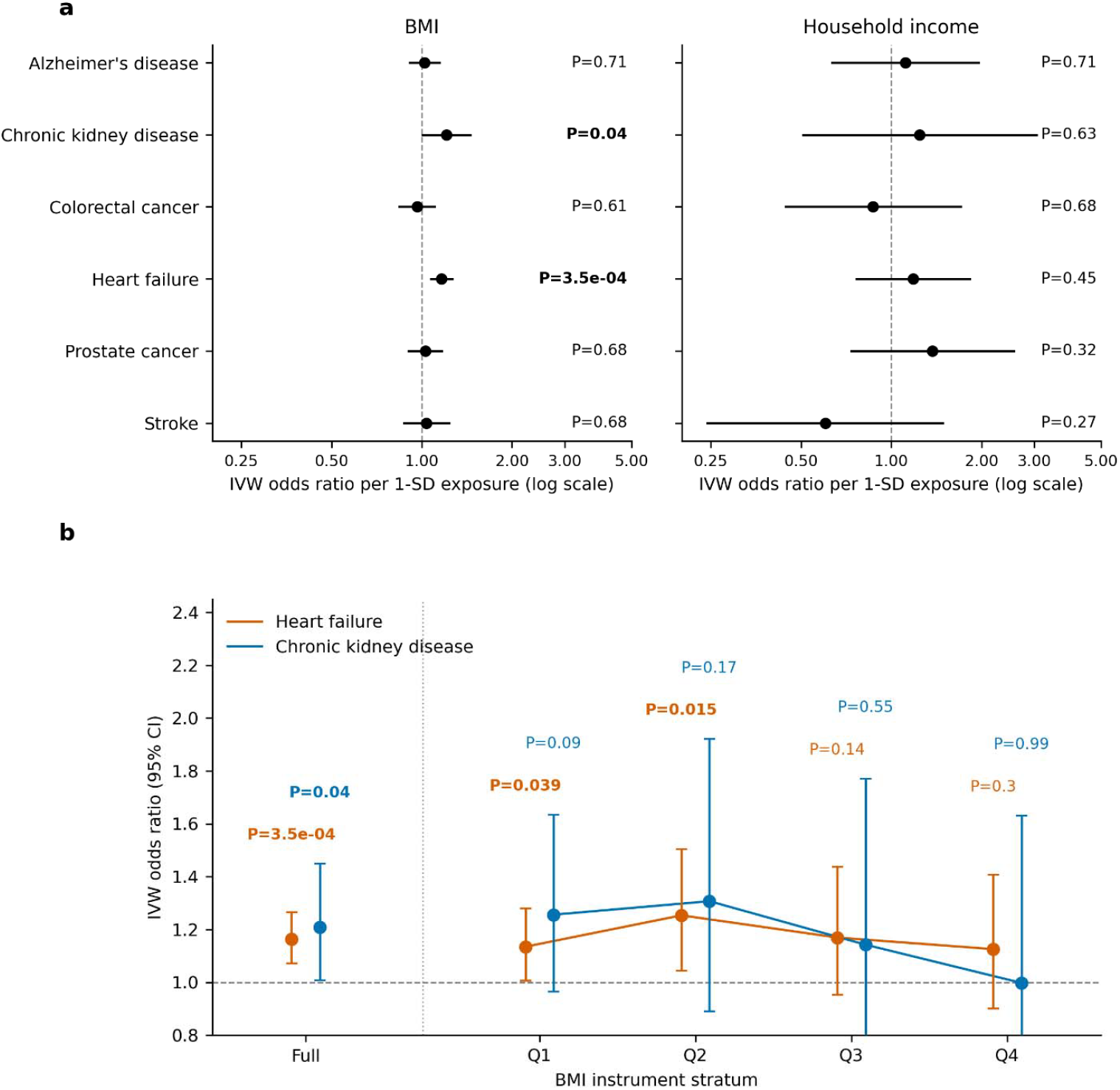
Positive-control Mendelian randomization against the Yang et al. disease-mortality GWAS. (a) Inverse-variance-weighted (IVW) odds ratio (OR) per 1-SD increase in the exposure for six disease-mortality outcomes, for body mass index (BMI) from Yengo et al.^2^ and for household income from Kweon et al.^6^. Horizontal lines are 95% confidence intervals; dashed vertical line marks OR = 1. IVW *P*-values are annotated per outcome and shown in bold for *P* < 0.05 associations; BMI analyses retained 779–877 variants and household-income analyses retained 32–38 variants across outcomes. (b) The BMI MR associations weaken in smaller, weaker instrument strata. IVW OR for BMI-heart failure (HF, orange) and BMI-chronic kidney disease (CKD, blue) mortality across the full BMI instrument set and four quartiles of those instruments stratified by BMI-exposure *P*-value, with Q1 the strongest and Q4 the weakest; a dotted vertical line separates the full set from the quartile subsets, and only Q1–Q4 are connected. BMI quartile strata retained 188–218 variants across outcomes after proxy search; per-stratum SNP counts are given in **Supplementary Table S1**. Error bars are 95% confidence intervals; *P*-values are annotated per point (bolded where *P* < 0.05).

Post-hoc power analyses also supported that, even with the full BMI instrument set, detectable effects varied substantially across outcomes (**Supplementary Table S2**). The minimum detectable OR at 80% power ranged from 1.13 for HF mortality to 1.30 for CKD mortality. Although power to detect an OR of 1.20 exceeded 80% for HF, AD, and prostate-cancer mortality, HF was the only outcome with both strong prior expectation for a BMI effect and clear empirical power. For household income, the minimum detectable OR exceeded 1.86 for all six outcomes and exceeded 2.50 for three outcomes. Thus, the analysis would have had limited ability to detect moderate associations, or even larger associations that remain plausible for weakly instrumented socio-economic traits.

As a positive control MR analysis, we next tested whether the same BMI instrument recovered expected associations when applied to case-control disease GWAS (**Figure 2a** and **Supplementary Table S3**). Genetically predicted BMI was associated with higher risk of HF (OR 1.66; 95% CI, 1.61–1.72; *P* = 2.9 × 10^−194^), ischaemic stroke (OR, 1.17; 95% CI, 1.12–1.22; *P* = 2.4 × 10^−12^), CKD (OR, 1.16; 95% CI, 1.10–1.23; *P* = 3.5 × 10^−7^) and colorectal cancer (OR, 1.12; 95% CI, 1.06–1.17; P = 7.5 × 10^−6^), and with lower risk of prostate cancer (OR, 0.87; 95% CI, 0.83–0.92; P = 6.3 × 10^−7^). The estimate for clinically diagnosed AD was not significant (OR, 0.88; 95% CI, 0.75–1.03; *P* = 0.12). Thus, the BMI instrument showed large and directionally coherent associations with disease risk for several outcomes, whereas corresponding evidence for disease mortality was limited. These results support the use of BMI as a positive-control exposure in our mortality MR analyses.

**Figure 2.**
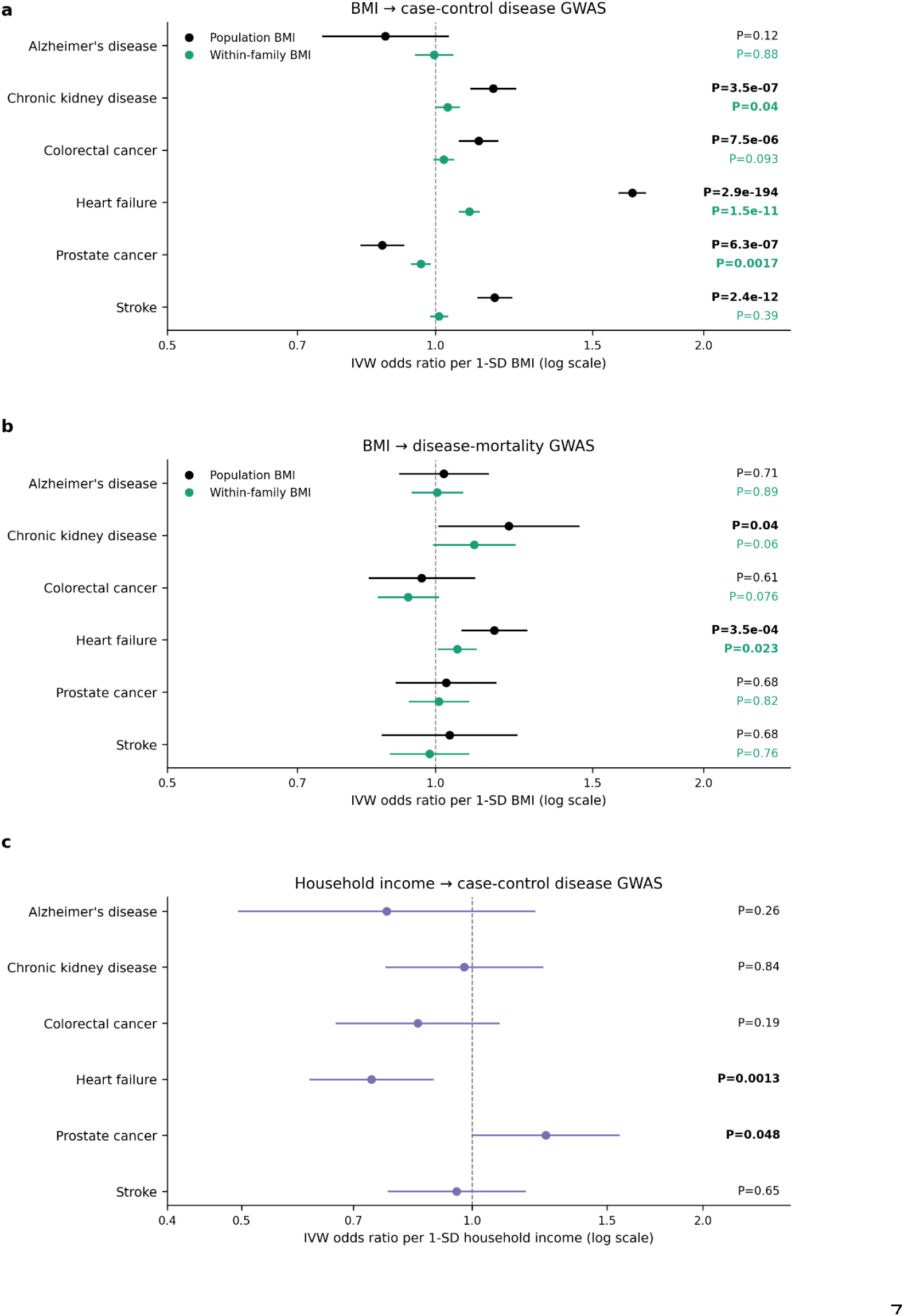
Additional positive-control validation and within-family sensitivity analyses. (a) Population-based BMI and within-family BMI against the six case-control disease GWAS. Points show inverse-variance-weighted (IVW) odds ratios (ORs) per 1-s.d. higher genetically predicted BMI for six disease-incidence outcomes: AD, CKD, colorectal cancer, HF, prostate cancer and ischaemic stroke. Horizontal lines denote 95% confidence intervals; the dashed vertical line marks OR = 1. Per-pair instrument counts and IVW *P* values are annotated, with bold *P* values denoting *P* < 0.05. (b) Population-based and within-family BMI against the six Yang et al.^1^ disease-mortality outcomes. Blue denotes population-based BMI instruments from Yengo et al.^2^; orange denotes within-family BMI instruments from Howe et al.^13^. Within-family BMI analyses retained 9–10 variants per outcome after harmonization and proxy search, whereas population-based BMI analyses retained 779–877 variants. Points show IVW ORs per 1-s.d. higher genetically predicted BMI; horizontal lines denote 95% confidence intervals; the dashed vertical line marks OR = 1. Per-pair instrument counts and IVW *P* values are annotated, with bold *P* values denoting *P* < 0.05. (c) Household income against the six case-control disease GWAS. Points show IVW ORs per 1-s.d. higher genetically predicted household income for the same six outcomes (32–38 variants per outcome). IVW *P* values are annotated per outcome with bold *P* values denoting *P* < 0.05.

The same positive-control effects were largely preserved when using within-family BMI as the exposure with 7–10 within-family instruments per outcome (**Figure 2a** and **Supplementary Table S3**). Within-family BMI was robustly associated with HF risk (OR 1.09; 95% CI, 1.06– 1.12; *P* = 1.5×10^−11^), prostate cancer (OR 0.96; 95% CI, 0.94–0.99; *P* = 1.7×10^−3^) and, at nominal significance, CKD (OR 1.03; 95% CI, 1.00–1.06; *P* = 0.04) suggesting that the associations are unlikely to be driven by dynastic or assortative-mating.

We ran a similar positive control MR using household income as the exposure (**Figure 2c** and **Supplementary Tables S3** and **S4**). Genetically proxied higher household income was associated with lower HF risk (OR, 0.74; 95% CI, 0.61–0.89; *P* = 1.3 × 10^−3^) and nominally associated with higher prostate cancer risk (OR, 1.25; 95% CI, 1.00–1.55; *P* = 0.048), whereas estimates for AD, CKD, colorectal cancer, and ischaemic stroke were not significant (all *P* > 0.18). This shows that even a weakly instrumented socio-economic exposure can detect associations in well-powered case-control GWAS, in contrast to the null mortality results, and consistent with limited effective power of the mortality GWAS.

We lastly repeated the BMI–mortality analysis using within-family BMI to assess whether the population-based BMI results were driven by demographic, assortative-mating or dynastic effects (**Figure 2b** and **Supplementary Tables S3** and **S4**). Using 9–10 within-family BMI instruments per outcome, genetically predicted BMI remained nominally associated with HF mortality (OR 1.06; 95% CI, 1.01–1.11; *P* = 0.02), whereas the CKD mortality estimate was attenuated and no longer conventionally significant (OR, 1.11; 95% CI, 1.00–1.23; *P* = 0.06). Estimates for AD, colorectal cancer, prostate cancer and stroke mortality were not significant (all *P* > 0.07). Although the HF and CKD point estimates were smaller than those obtained using population-based BMI instruments, confidence intervals overlapped across all six outcomes, indicating no qualitative change in inference.

## Discussion

Our results indicate that disease mortality GWAS can currently detect only relatively large MR effects, and primarily when the exposure is well instrumented. Even using BMI, a highly heritable and polygenic trait as a positive control, we detected an association in only two of six disease mortality outcomes pertaining to HF and CKD. Stratifying the BMI genetic instruments into smaller, weaker subsets attenuated the HF association and eliminated the CKD association. Our results do not refute the possibility that susceptibility and survival have partly distinct genetic architectures; rather, they show that the current mortality GWAS are likely not yet sufficiently powered to fully answer this question.

This analysis has limitations. Chiefly, MR against disease-specific mortality endpoints does not isolate post-diagnosis survival from disease ascertainment, severity, treatment, and competing-risk processes; therefore, our estimates may reflect a mixture of effects on disease incidence, progression, and survival after diagnosis. Conclusions about limited susceptibility-survival overlap should therefore be interpreted as provisional until mortality GWAS are sufficiently powered, or explicitly calibrated, to detect shared effects if they exist. To conclude, more research and larger sample sizes are required to draw stronger conclusions on the value of prognosis GWAS.

## Ethics approval and consent to participate

This commentary used only publicly available GWAS summary statistics. Ethics approvals for the underlying GWAS are described in the original publications.

## Acknowledgements

Chen-Yang Su is supported by a Postdoctoral Research Scholarship (https://doi.org/10.69777/2005819) from the Fonds de recherche du Québec. The funders had no role in the conceptualization, study design, data collection, analysis, decision to publish, or preparation of the manuscript. Guillaume Butler-Laporte receives salary support from the FRQS. We thank the authors of Yang et al. and the participants of the contributing biobanks whose data made this commentary possible, and the investigators of the GIANT, UK Biobank and household-income GWAS consortia for making their summary statistics publicly available.

## Data availability

All summary statistics are publicly available and described in **Supplementary Table S5**.

Yang et al. disease-mortality GWAS: https://figshare.com/projects/Progression_GWAS/252002.

BMI (Yengo et al.): https://portals.broadinstitute.org/collaboration/giant/.

Household income (Kweon et al.): https://figshare.com/projects/Progression_GWAS/252002.

Within-family BMI (Howe et al.): https://opengwas.io/datasets/ieu-b-4815

The disease-diagnosis GWAS used for positive-control validation were publicly available: Kunkle et al. 2019 for AD through NIAGADS NG00075; Wuttke et al. 2019 CKDGen Round 4 European-ancestry GWAS for CKD; Fernandez-Rozadilla et al. 2023 for colorectal cancer, Wang et al. 2023 European-ancestry GWAS for prostate cancer and Mishra et al. 2022 GIGASTROKE European-ancestry ischaemic stroke GWAS through the GWAS Catalog; and Henry et al. 2025 HF GWAS through the HERMES consortium.

## Code availability

We used R v4.1.2, TwoSampleMR v0.5.6, gwasvcf, PLINK v1.9. All MR analyses adhered to the STROBE-MR guidelines^14^ (**Supplementary Note 1**).

## Author contributions

**Chen-Yang Su:** Data curation; formal analysis; methodology; software; validation; visualization; writing – original draft; writing – review and editing.

**Guillaume Butler-Laporte**: Conceptualization; funding acquisition; supervision; writing – review and editing.

